# Point-of-Care Ultrasound as a Prognostic Tool in Critically Ill Patients: Insights Beyond Core Muscle Mass

**DOI:** 10.1101/2025.03.19.25324253

**Authors:** Rachel Skoczynski, Jonathan Hansen, Sanjib Das Adhikary, Erik Lehman, Anthony S Bonavia

## Abstract

**Background:** Muscle wasting is a major concern in ICUs, contributing to morbidity, mortality, and prolonged rehabilitation. While CT-derived L3 Skeletal Muscle Index (L3SMI) assesses core muscle mass, it may not capture peripheral muscle atrophy or fluid-based changes. Point-of-care ultrasound (POCUS) offers a rapid, non-invasive alternative. This study evaluated the prognostic value of POCUS-based muscle measurements compared with L3SMI in predicting mortality, frailty, and functional outcomes.

**Methods:** In this prospective study, 50 critically ill adults meeting Sepsis-3 criteria or requiring respiratory/vasopressor support underwent POCUS assessments of biceps brachii, rectus femoris, and vastus intermedius thickness at days 1, 7, and 14 post-ICU admission. Twenty-eight patients also had CT scans within seven days for L3SMI calculation. The primary outcome was 90-day mortality; secondary outcomes included in-hospital and 30-day mortality, Clinical Frailty Score, and Zubrod/ECOG performance status. Muscle measurements were analyzed both raw and indexed to body surface area, with predictive performance assessed via correlation and ROC analysis.

**Results:** Day 1 biceps brachii thickness strongly predicted in-hospital mortality (AUC 0.84; sensitivity 1.0, specificity 0.67) and retained predictive value for 30-day and 90-day mortality. Vastus intermedius thickness on day 1 was moderately predictive (AUC 0.79). At later time points, larger vastus intermedius measurements correlated negatively with ICU-and ventilator-free days, suggesting edema-related pseudohypertrophy. L3SMI did not significantly correlate with ultrasound-based muscle measurements or clinical outcomes. POCUS-derived peripheral muscle indexing was associated with frailty indices, highlighting its role in capturing meaningful functional deficits.

**Conclusion:** POCUS-based muscle assessments, particularly of the biceps brachii and vastus intermedius, provide valuable prognostic insights beyond conventional L3SMI. While L3SMI remains a core muscle measure, fluid shifts and localized muscle wasting in critical illness may be better captured by ultrasound.

## Introduction

In critically ill patients, timely and accurate prognostication is essential for guiding clinical decisions and improving patient outcomes. While various tools have been utilized to assess the severity of illness and predict recovery, point-of-care ultrasound (POCUS) serves as a rapidly accessible and non-invasive method to provide valuable insight to real-time patient assessment. Traditionally, ultrasound has been used in monitoring hemodynamic, cardiac function, and fluid status.^1^ More recently, its role in evaluating skeletal muscle mass has emerged as an area of interest given that muscle wasting is known to have profound implications in critically ill adults. ^2^

During critical illness, muscle mass can decrease by as much as 15% with one week, which equates to nearly 2% of skeletal muscle loss per day during the first week of ICU admission.^3, 4^ The accelerated rate of muscle mass loss in critically ill patients stems from multiple converging processes, including upregulated catabolic pathways, suppressed protein synthesis, excess cortisol, and reduced physical activity, all of which contribute to muscle atrophy. ^5^ Furthermore, low muscle mass is associated with a myriad of negative health outcomes: prolonged hospitalization, hospital readmission, increased risk of long-term disability, increased risk of mortality, and poor functional recovery.^6–8^

Computed tomography (CT) has traditionally been a central diagnostic tool in critical care and is often used to quantify muscle loss by focusing on core musculature. A key metric, the L3 Skeletal Muscle Index (L3SMI), involves measuring the cross-sectional area of core muscles—such as the psoas major, erector spinae, quadratus lumborum, paraspinals, transversus abdominis, rectus abdominis, and internal and external obliques—at the third lumbar vertebra (L3) and normalizing this area to body surface area.^9^ Although L3SMI is a popular method for assessing muscle mass, low values can be mistaken for sarcopenia unless reduced muscle function is also demonstrated.^10^

However, relying solely on core muscle measurements may overlook key aspects of a patient’s condition, particularly fluid imbalances arising from systemic inflammation, organ failure, and intravenous fluid administration. Such fluid shifts can distort muscle appearance on imaging, making tissues appear either edematous or atrophied regardless of true muscle mass.^11^ By contrast, point-of-care ultrasound (POCUS) can rapidly evaluate peripheral muscle and thus offer a more comprehensive view of patient status. Incorporating POCUS findings into clinical practice may facilitate more accurate predictions of prognosis and guide targeted rehabilitation strategies in critically ill populations.^12^

The goal of the present study was to explore the utility of POCUS as prognostic tool in critically ill patients. We hypothesized that appendicular muscle mass, assessed using POCUS, would correlate with non-appendicular muscle mass, quantified by L3SMI, and that both measures would predict long-term outcomes following hospitalization of critically ill patients. Our primary clinical outcome was 90-day mortality, and secondary outcome was Clinical Frailty Scores.

## Results

### Patient Cohort

Fifty patients were included in the investigation, with a mean age of 62.9 ± 16.6 years, mean APACHE II score of 21.2 ± 7.2, and mean SOFA score 8.3 ± 3.9 on the day of enrollment (**Table 1**). Recruitment and follow-up data is illustrated in **Supplementary Figure S1**. Clinical outcomes for these critically ill patients are shown in **Table 2**.

**Table 1.** Demographics and Patient Characteristics.

| Metric | Units | Ultrasound Only<br>(n=22) | Ultrasound and<br>Abdominal<br>Computed<br>Tomography (CT)<br>scan (n=28) | All Patients (n=50) |
| --- | --- | --- | --- | --- |
| Age | years | 62.7 ± 14.0 | 63.1 ± 18.7 | 62.9 ± 16.6 |
| Body surface area | m<sup>2</sup> | 2.09 ± 0.36 | 2.02 ± 0.31 | 2.05 ± 0.33 |
| APACHE II score |  | 19.2 ± 6.0 | 22.7 ± 7.9 | 21.2 ± 7.2 |
| SOFA score, day 1 of<br>critical illness |  | 7.3 ± 3.8 | 9.2 ± 3.9 | 8.3 ± 3.9 |
| Body Mass Index | kg/m<sup>2</sup> | 32.3 ± 10.1 | 30.5 ± 10.1 | 31.3 ± 10.0 |
| Lactate | mmol/L | 3.1 ± 2.1 | 5.9 ± 6.7 | 4.7 ± 5.4 |
| Bicarbonate | mmol/L | 20.3 ± 5.5 | 18.5 ± 4.5 | 19.3 ± 5.0 |
| Leukocyte count | x10<sup>9</sup>/L | 13.6 ± 8.2 | 20.0 ± 9.0 | 17.2 ± 9.2 |
| C-reactive protein | mg/L | 12.3 ± 12.2 | 17.7 ± 11.0 | 15.4 ± 11.6 |
| Creatinine | mg/dL | 2.2 ± 2.2 | 2.0 ± 1.5 | 2.1 ± 1.8 |
| Hematocrit | % | 32.1 ± 5.5 | 30.7 ± 5.5 | 31.3 ± 5.5 |
| Procalcitonin | ng/mL | 0.4 ± 0.5 | 8.3 ± 7.8 | 6.3 ± 7.5 |
| Albumin | g/dL | 3.0 ± 0.5 | 2.7 ± 0.7 | 2.8 ± 0.6 |
| Protein | g/dL | 5.3 ± 1.5 | 5.8 ± 1.2 | 5.6 ± 1.4 |
| Total bilirubin | mg/dL | 1.9 ± 3.67 | 2.2 ± 2.3 | 2.0 ± 3.0 |
| Sepsis |  | 8 (36.4%) | 16 (57.1%) | 24 (48.0%) |
| Shock |  | 7 (31.8%) | 15 (53.6%) | 22 (44.0%) |
| Acute Kidney Injury |  | 5.0 (22.7%) | 12.0 (42.9%) | 17.0 (34.0%) |
| On Vasopressor<br>infusion |  | 13 (59.1%) | 24 (85.7%) | 37 (74.0%) |

**Table 2.** Clinical Outcomes.

|  | Ultrasound Only<br>(n=22) | Ultrasound and<br>Abdominal<br>Computed<br>Tomography (CT)<br>scan (n=28) | All Patients<br>(n=50) |
| --- | --- | --- | --- |
| **In-hospital mortality** | 2 (9.1%) | 4 (14.3%) | 6 (12.0%) |
| **30-day mortality** | 4 (18.2%) | 7 (25.0%) | 11 (22.0%) |
| **90-day mortality** | 6.0 (27.3%) | 10.0 (35.7%) | 16.0 (32.0%) |
| **90-day hospital readmission** | 1 (4.5%) | 4 (14.3%) | 5 (10.0%) |
| **28-day ventilator-free days** | 24.23 ± 6.06 | 22.39 ± 9.64 | 23.20 ± 8.24 |
| **28-day hemodialysis-free days** | 23.18 ± 9.67 | 22.07 ± 11.24 | 22.56 ± 10.49 |
| **28-day vasopressor-free days** | 25.14 ± 5.77 | 22.11 ± 9.25 | 23.44 ± 7.99 |
| **28-day ICU-free days** | 20.00 ± 7.55 | 18.39 ± 9.69 | 19.10 ± 8.77 |
| **28-day hospital-free days** | 14.05 ± 9.15 | 12.29 ± 10.11 | 13.06 ± 9.64 |
| **28-day organ dysfunction-free days** | 21.32 ± 9.14 | 21.54 ± 9.24 | 21.44 ± 9.10 |
| **28-day Clinical Frailty Score** | 4.77 ± 3.04 | 5.54 ± 2.94 | 5.20 ± 2.98 |
| **Discharge Location** |  |  |  |
| ---In-hospital death | 0 | 4 | 4 |
| ---Long-term acute care hospital | 3 | 1 | 4 |
| ---Skilled nursing facility or rehabilitation facility | 9 | 12 | 21 |
| ---Home | 9 | 10 | 19 |
| ---Hospice | 1 | 1 | 2 |
| **Residence at 30 Days** |  |  |  |
| ---Home | 10 | 11 | 21 |
| ---Assisted Living Facility | 4 | 1 | 5 |
| ---Hospital | 2 | 6 | 8 |
| **Zubrod/ECOG Score at 30 days** |  |  |  |
| --- 0: Fully active, able to carry on all pre-disease performance without restriction (normal activity). | 4 | 4 | 8 |
| --- 1: Restricted in physically strenuous activity but ambulatory and able to carry out work of a light or sedentary nature. | 5 | 3 | 8 |
| --- 2: Ambulatory and capable of all self-care but unable to carry out any work activities; up and about more than 50% of waking hours. | 0 | 2 | 2 |
| **--- 3: Capable of only limited self-care; confined to bed or chair more than 50% of waking hours.** | **2** | **5** | **7** |
| **--- 4: Completely disabled; cannot carry out any self-care; totally confined to bed or chair.** | **5** | **3** | **8** |
| **--- 5: Deceased** | **2** | **4** | **6** |

**Table 3.** Core and Peripheral Muscle Measurements.

|  | Units | Ultrasound Only (n=22) | Ultrasound and Abdominal Computed Tomography (CT) scan (n=28) | All Patients (n=50) |
| --- | --- | --- | --- | --- |
| **Skeletal Muscle Area, by CT scan, at L3 level** | cm<sup>2</sup> | N/A |  |  |
| --Males |  |  | 145.3 ± 30.8 (n=19) |  |
| --Females |  |  | 103.3 ± 25.3 (n=9) |  |
| **Skeletal Muscle Index, by CT scan, at L3 level** | cm<sup>2</sup>/m<sup>2</sup> | N/A |  |  |
| --Males |  |  | 47.4 ± 10.2 (n=19) |  |
| --Females |  |  | 39.4 ± 9.43 (n=9) |  |
| **Low muscle mass, by L3SMI criteria** |  | N/A | 7 (n=19) |  |
| **Mean Biceps brachii, day 1** | cm | 1.73 ± 0.58 | 1.67 ± 0.60 | 1.70 ± 0.58 |
| **Biceps brachii indexed to BSA, day 1** | cm/m<sup>2</sup> | 0.83 ± 0.24 | 0.82 ± 0.24 | 0.82 ± 0.24 |
| **Mean Rectus femoris, day 1** | cm | 1.15 ± 0.65 | 0.96 ± 0.55 | 1.04 ± 0.60 |
| **Mean Rectus femoris indexed to BSA, day 1** | cm/m<sup>2</sup> | 0.53 ± 0.24 | 0.47 ± 0.26 | 0.50 ± 0.25 |
| **Mean Vastus intermedius, day 1** | cm | 1.06 ± 0.48 | 0.93 ± 0.53 | 0.99 ± 0.51 |
| **Vastus intermedius indexed to BSA, day 1** | cm/m<sup>2</sup> | 0.51 ± 0.19 | 0.46 ± 0.23 | 0.48 ± 0.21 |
| **Mean Biceps brachii, day 7** | cm | 1.95 ± 0.92 | 1.70 ± 0.82 | 1.80 ± 0.85 |
| **Mean Biceps brachii indexed to BSA, day 7** | cm/m<sup>2</sup> | 0.94 ± 0.36 | 0.79 ± 0.28 | 0.85 ± 0.32 |
| **Mean Rectus femoris, day 7** | cm | 1.19 ± 0.65 | 0.88 ± 0.38 | 1.01 ± 0.53 |
| **Mean Rectus femoris indexed to BSA, day 7** | cm/m<sup>2</sup> | 0.56 ± 0.21 | 0.42 ± 0.16 | 0.48 ± 0.19 |
| **Mean Vastus intermedius, day 7** | cm | 1.04 ± 0.49 | 0.86 ± 0.44 | 0.94 ± 0.46 |
| **Mean Vastus intermedius indexed to BSA, day 7** | cm/m<sup>2</sup> | 0.49 ± 0.14 | 0.40 ± 0.17 | 0.44 ± 0.16 |
| **Mean Biceps brachii, day 14** | cm | 1.21 ± 0.32 | 1.91 ± 1.01 | 1.52 ± 0.75 |
| **Mean Biceps brachii indexed to BSA, day 14** | cm/m<sup>2</sup> | 0.63 ± 0.21 | 0.90 ± 0.45 | 0.75 ± 0.34 |
| **Mean Rectus femoris, day 14** | cm | 0.89 ± 0.22 | 1.24 ± 0.62 | 1.05 ± 0.45 |
| **Mean Rectus femoris indexed to BSA, day 14** | cm/m<sup>2</sup> | 0.45 ± 0.07 | 0.59 ± 0.33 | 0.51 ± 0.22 |
| **Mean Vastus intermedius, day 14** | cm | 0.78 ± 0.12 | 0.96 ± 0.35 | 0.86 ± 0.25 |
| **Mean Vastus intermedius indexed to BSA, day 14** | cm/m<sup>2</sup> | 0.40 ± 0.07 | 0.46 ± 0.20 | 0.43 ± 0.13 |

\*BSA: body surface area, calculated by Moeller formula

Briefly, nearly half (48%) of the cohort met sepsis criteria, and 44% experienced shock, while 74% required vasopressor support. Renal impairment was common, with 34% developing acute kidney injury. Mean BMI was 31.3 ± 10.0, and average laboratory values (e.g., lactate 4.7 ± 5.4 mmol/L, leukocyte count 17.2 ± 9.2 × 10^9^/L, and procalcitonin 6.3 ± 7.5 ng/mL) reflected the high acuity of these patients.

Clinical outcomes were similarly indicative of severe illness. In-hospital mortality was 12%, rising to 22% at 30 days and 32% at 90 days, and 10% of survivors were readmitted within 90 days. Participants averaged 23.2 ± 8.2 ventilator-free days and 19.1 ± 8.8 ICU-free days over the first 28 days, although many required ongoing care post-discharge: 42% were transferred to a skilled nursing or rehabilitation facility, and only 38% were discharged directly home. At 30 days, 21 patients resided at home, while 13 remained in hospital or required assisted living, highlighting the long-term impact of critical illness on functional recovery.

### Skeletal Muscle Measurements

Core muscle measurements, measured by abdominal CT imaging, and peripheral muscle measurements, measured by point-of-care ultrasound, are shown in **Table 2** and **Fig 1**. Twenty-two patients underwent only ultrasound assessment of muscle mass, while 28 patients underwent both ultrasound and CT scan within 7 days of enrollment.

**Fig 1.**
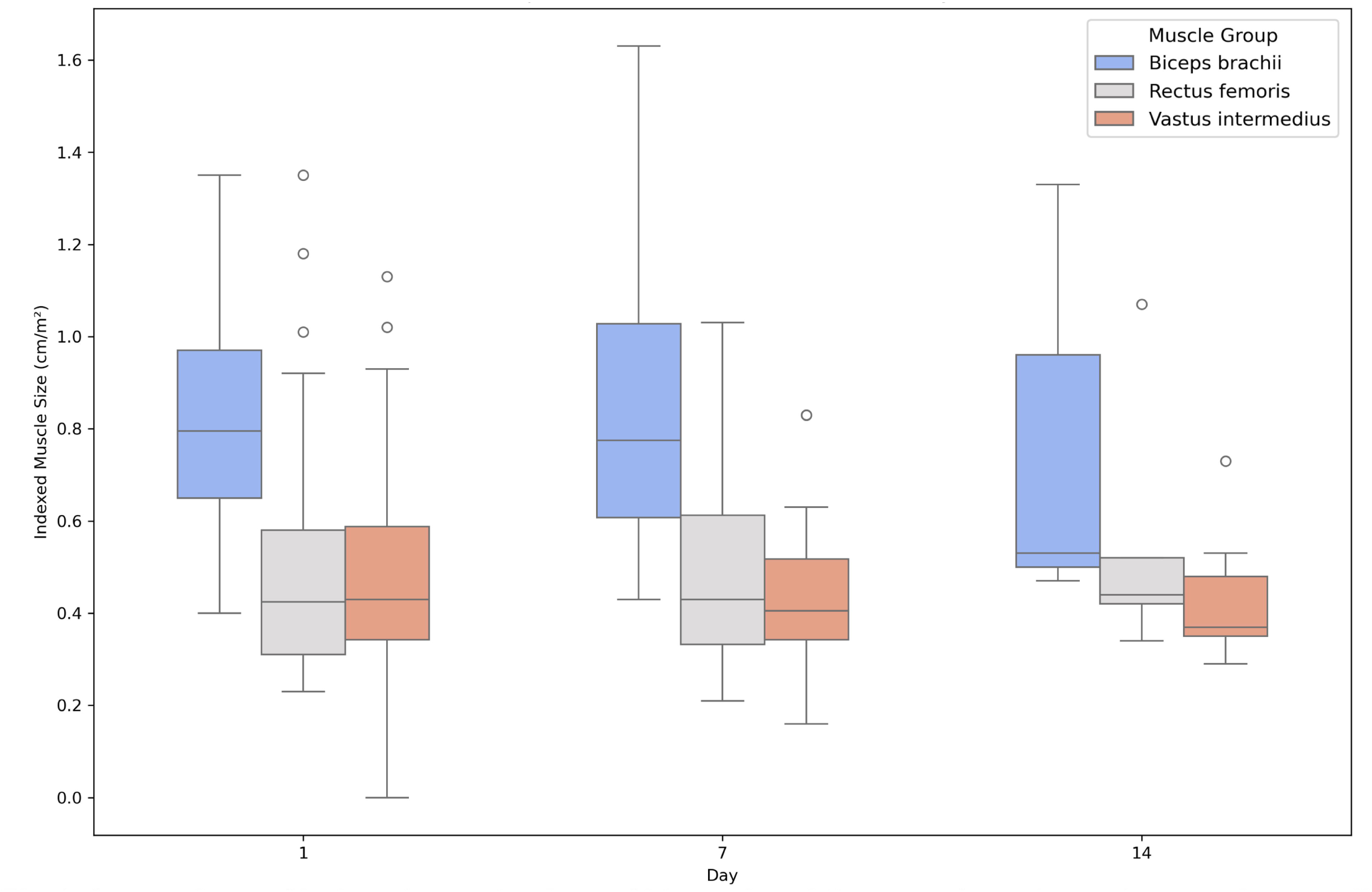
Comparison of indexed muscle sizes of biceps brachii, rectus femoris, and vastus intermedius at days 1, 7, and 14 following onset of critical illness. Boxplots depict the distribution of muscle sizes indexed to body surface area for each muscle group on the specified days, in cm/m^2^. The median and interquartile range are shown within each box, with whiskers extending to 1.5 times the interquartile range. Outliers are represented as individual points. n=50 at day 1, n=26 at day 7, and n=9 at day 14.

### L3 Skeletal Muscle Index Does Not Correlate with Peripheral Muscle Wasting

Our analysis revealed no significant correlation between L3SMI and peripheral muscle measurements obtained via ultrasound, including the biceps brachii, rectus femoris, and vastus intermedius sizes. Statistically, the correlation coefficients between L3SMI and peripheral muscles of the upper and lower extremities were consistently weak and not significant (e.g., biceps brachii: Spearman r = −0.12, *P* = 0.36; rectus femoris: r = 0.10, *P* = 0.42; vastus intermedius: r = −0.08, *P* = 0.51). Furthermore, L3SMI was not predictive of frailty as quantified by Clinical Frailty Score (CFS) or Zubrod score, nor did it correlate with ICU-free days, ventilator-free days, or vasopressor-free days.

### Biceps Brachii as a Strong Predictor of In-Hospital Mortality

Among muscle measurements, the biceps brachii size and BSA-indexed biceps brachii size on day 1 (Bb_d1_ and Bb_d1i_, respectively) emerged as robust predictors of the primary outcome, in-hospital mortality (**Fig 2**). Bb_d1_ demonstrated the highest predictive accuracy with an AUC of 0.84 (95% CI: 0.69–0.99, *P* < 0.001), perfect sensitivity (1.0), and a specificity of 0.67. When indexed to BSA, the predictive performance of Bb_d1i_ remained notable, achieving an AUC of 0.74 (95% CI: 0.48–1.00, *P =*0.071) with improved specificity (0.91) but reduced sensitivity (0.6). Similarly, vastus intermedius size on day 1 (Vi_d1_) achieved statistical significance, with an AUC of 0.79 (95% CI: 0.63– 0.95, *P* < 0.001), sensitivity of 1.0, and specificity of 0.6, while its BSA-indexed counterpart (Vi_d1i_) had an AUC of 0.73 (95% CI: 0.54–0.92, *P* = 0.016), sensitivity of 1.0, and specificity of 0.49.

**Fig 2.**
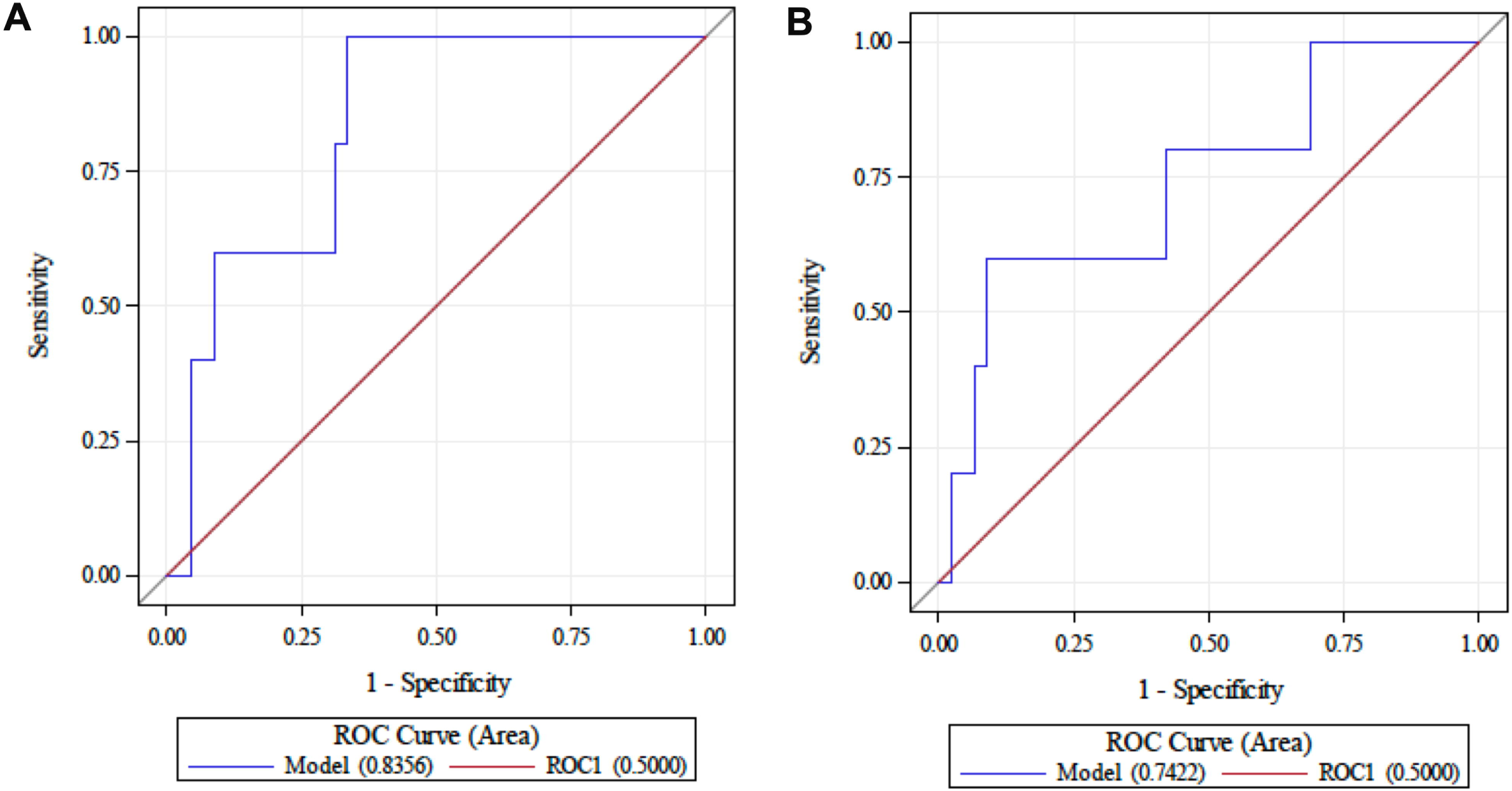
Biceps brachii size as a predictor of in-hospital mortality, in critically ill patients. (A) Biceps brachii, measured on day 1, demonstrates AUC 0.84 (95% Cl 0.69 - 0.99, p<0.001), Youden index= 0.67, mean size= 1.57 cm, sensitivity= 1, specificity= 0.67. (B) Biceps brachii, measured on day 1 and indexed to body surface area, demonstrates AUC 0.74 (95% Cl 0.48-1.00, p=0.071, n=50), Youden index= 0.51, mean= 0.55 cm/m2, sensitivity= 0.6, specificity= 0.91. n=50.

**Fig 3.**
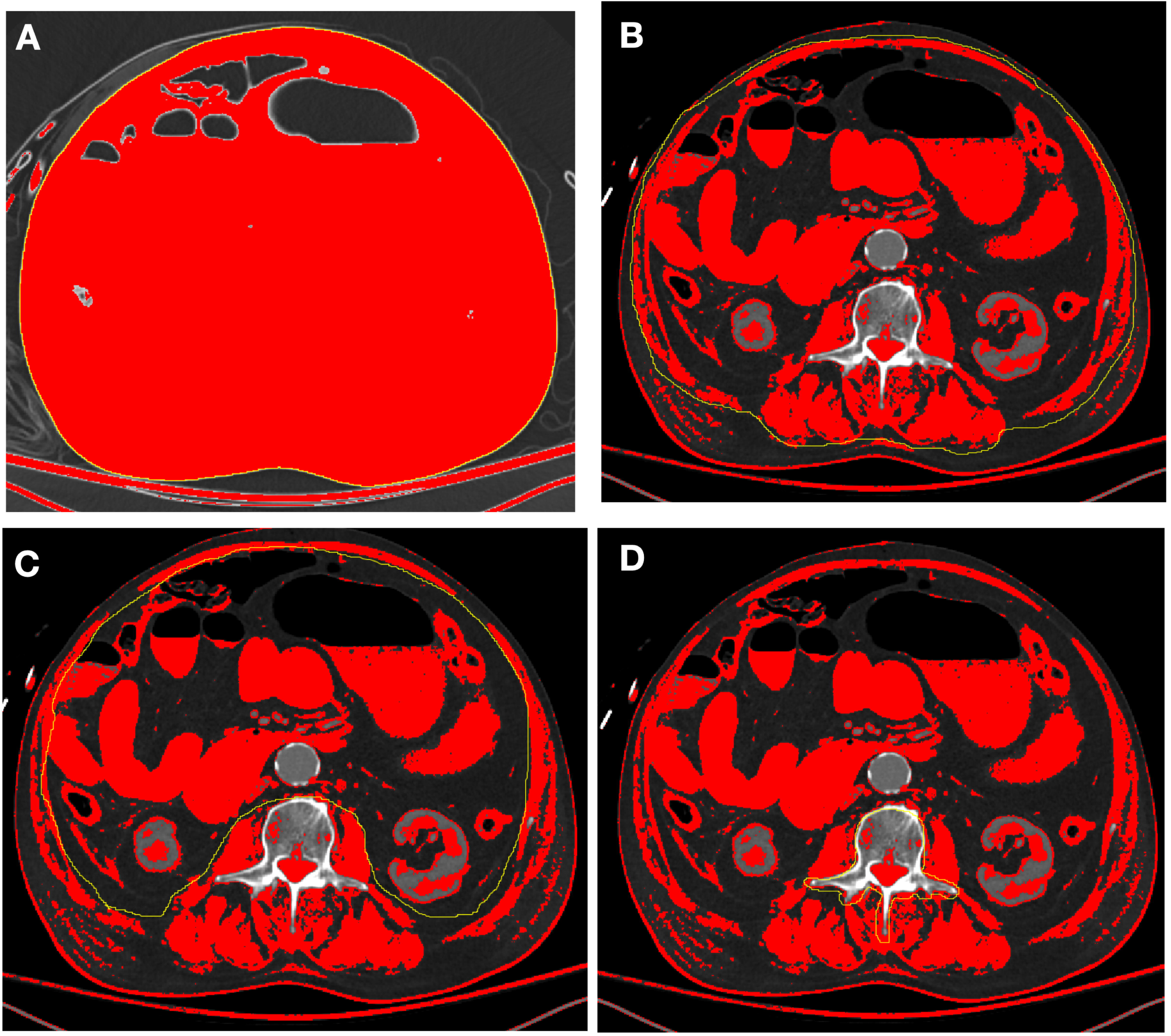
Quantification of Skeletal Muscle Area at the L3 Axial Level Using Computed Tomography (CT) and lmageJ Software. (A) The entire abdominal wall is manually traced (yellow outline) to establish the total region of interest. (B) The outer boundary of the abdominal muscles is delineated (yellow outline), and the corresponding area is calculated. (C) The inner boundary of the abdominal muscles is then traced (yellow outline), and its area is measured. (D) The vertebral region is excluded by subtracting its area. To distinguish skeletal muscle, a radiodensity threshold range of-29 to 150 Hounsfield units (HU) was applied.

**Fig 4.**
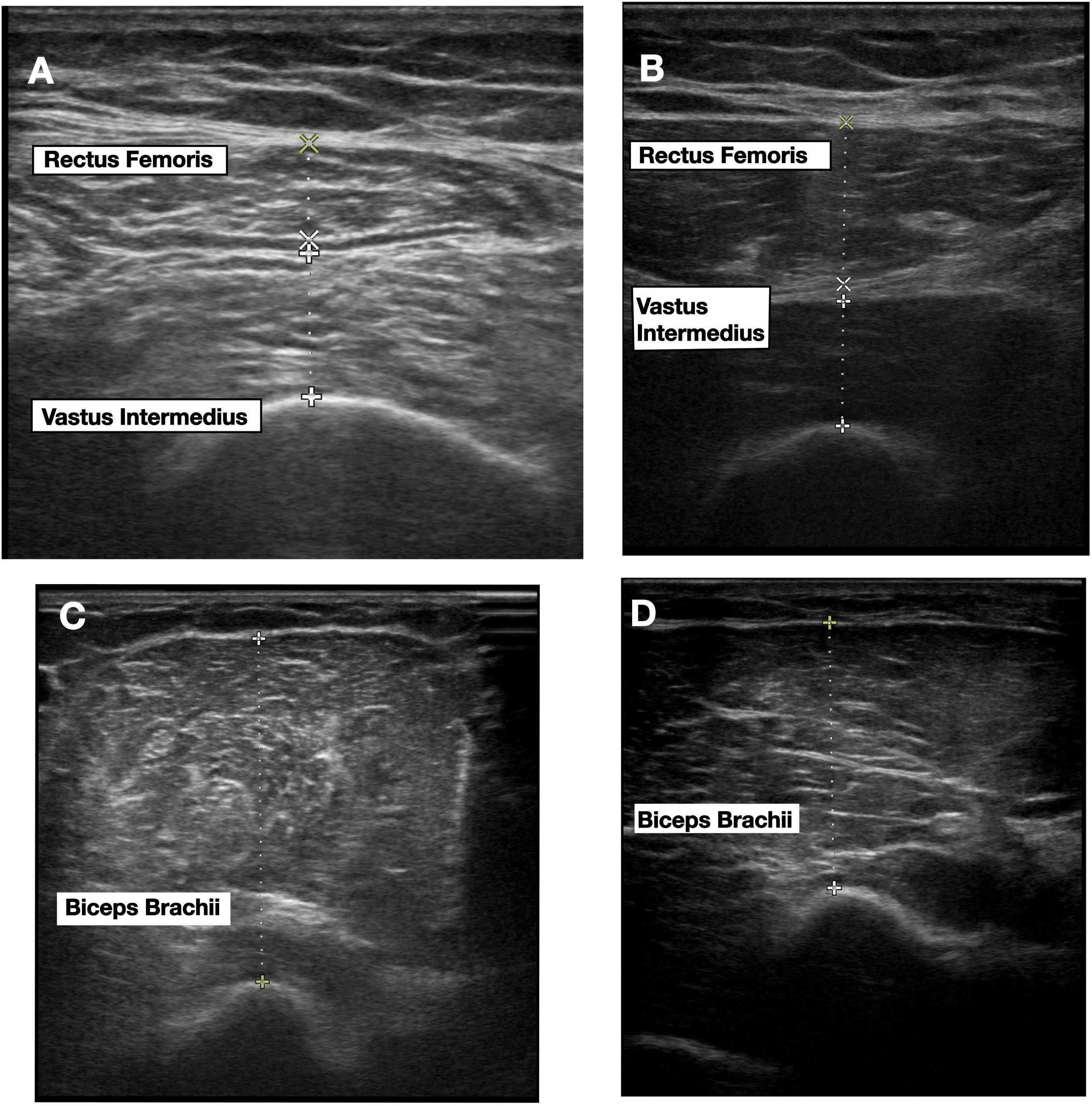
Point-of-care Ultrasound for Peripheral Muscle Measurement. Images (A) and (B) demonstrate measurements of rectus femoris and vastus intermedius muscles from two different patients, while images (C) and (D) demonstrate measurement of biceps brachii in these same two patients.

### Biceps Brachii Indices as Predictors of 30-Day and 90-Day Mortality

For 30-day mortality, the biceps brachii size on day 1 (Bb_d1_) again demonstrated the highest predictive accuracy, with an AUC of 0.83 (95% CI: 0.70–0.96, *P* < 0.001), sensitivity of 1.0, and specificity of 0.68, maintaining its robustness when indexed to BSA (AUC = 0.83, 95% CI: 0.51–0.95, *P* = 0.039). For 90-day mortality, Bb_d1_ also performed strongly, achieving an AUC of 0.76 (95% CI: 0.62–0.89, *P* < 0.001), sensitivity of 1.0, and specificity of 0.54. Measurements at later time points were also informative; Bb_d7i_ (day 7, indexed to body surface area) achieved an AUC of 0.77 (95% CI: 0.50–1.00, *P* = 0.051), while Bb_d14i_ (day 14, indexed) displayed the highest overall accuracy for 90-day mortality, with an AUC of 0.88 (95% CI: 0.63–1.00, *P* = 0.003).

Vastus intermedius (Vi_d1_) had weaker, though still significant, predictive power for both 30-day (AUC = 0.71, *P* = 0.054) and 90-day mortality (AUC = 0.69, *P* = 0.023). These findings reinforce the superior utility of biceps brachii indices, particularly the indexed values at later time points, for predicting both short-and long-term mortality.

### Relationship Between Muscle Size and Frailty Scores

Indexed measurements of muscle size, including the biceps brachii, rectus femoris, and vastus intermedius on day 1 of illness, were found to significantly correlate with and predict frailty, as assessed by the Clinical Frailty Score (CFS) and Zubrod/ECOG score. Each 1 cm/m^2^ increase in indexed biceps size was associated with a 4.5-point decrease in CFS, while each 1 cm/m^2^ increase in indexed vastus or rectus size was associated with a 3.9-point decrease in CFS. Weak but significant negative correlations were observed between day 1-indexed biceps brachii (Spearman’s ρ =-0.31, *P* = 0.03), rectus femoris (ρ =-0.34, *P* = 0.003), and vastus intermedius (ρ =-0.32, *P* = 0.024) with the CFS.

Biceps index was also significantly correlated with 30-day Zubrod score (ρ = - 0.42, *P* = 0.036), with each 1 cm/m^2^ increase in biceps index being associated with 4.2-point decrease in Zubrod score by quantile regression analysis (*P* = 0.036). Indexed rectus measurements at day 1 were also strongly predictive of 30-day function (each 1 cm/m^2^ increase in muscle size predicting a 3.4-point decrease in Zubrod score, p=0.023), as were indexed vastus measurements (1 cm/m^2^ increase in muscle size predicting a 4.1-point decrease in Zubrod score, *P* = 0.004). Given that the Zubrod scale ranges from one to five, these effects of peripheral muscle size on function are significant.

In contrast, low muscle mass as determined by the L3SMI did not correlate with frailty, as assessed by CFS or Zubrod Performance Status, highlighting the importance of localized muscle size measurements in assessing functional outcomes.

### Relationship Between Muscle Measurements at Later Hospitalization Stages and Clinical Outcomes

Indexed muscle measurements at later stages of hospitalization were consistently negatively correlated with critical illness outcomes, suggesting that tissue edema rather than true hypertrophy may account for observed increases in size. For instance, indexed biceps brachii size on Day 14 showed a strong negative correlation with 28-day ICU-free days (Spearman r = –0.81, *P =* 0.004). Similarly, biceps size and 28-day ventilator-free days shifted from a weak positive correlation on Day 1 (r = 0.32) to a weak negative correlation on Day 14 (r = –0.35). The vastus intermedius exhibited a comparable pattern: on Day 14, its indexed thickness demonstrated a moderate negative correlation with ventilator-free days (r = –0.46) and a strong negative correlation with 28-day organ-dysfunction-free days (r = –0.76, *P =* 0.010), persisting even when normalized to body surface area (r = –0.56, *P =* 0.102). Additionally, changes in vastus size from Days 7 to 14 were moderately negatively correlated with multiple outcomes (r = –0.61, *P =* 0.067), underscoring how fluid retention and inflammatory processes can inflate ultrasound measurements and obscure actual muscle status.

## Discussion

This study examined the size of various muscle groups (biceps brachii, vastus intermedius, and rectus femoris) in critically ill adults, revealing that each muscle group provided distinct and valuable insights into clinical outcomes. These findings underscore the importance of assessing multiple muscle groups to enhance prognostication and guide clinical decision-making in critical care.

In the present analysis, the biceps brachii size measured early during critical illness demonstrated strong predictive capabilities for short-and long-term mortality outcomes. Its size on day 1 showed significant predictive power for in-hospital mortality, indicating its utility as an early marker of systemic inflammation and catabolic states in critically ill patients. This aligns with prior studies linking upper limb muscle atrophy to adverse outcomes due to heightened proteolysis during systemic inflammation.^13^ However, the predictive strength of the biceps brachii diminished at later time points, possibly reflecting the effects of fluid shifts or compensatory hypertrophy during recovery.^14^

Although a larger vastus intermedius muscle might intuitively suggest better function, our data at later hospitalization stages indicate that greater size on Day 14 is actually negatively correlated with 28-day ICU-free days, organ-dysfunction-free days, and ventilator-free days, and predicts higher 90-day readmission rates. This initially appears paradoxical, yet the most likely explanation is fluid retention and edema rather than true muscular hypertrophy. Critically ill patients often accumulate fluid over time, which can artifactually enlarge muscle dimensions on ultrasound. Several studies have documented substantial fluid accumulation in critically ill patients and those with sepsis, over the course of an ICU stay, often amounting to 5-10 liters of net positive fluid balance in the first one to two weeks of critical illness.^15–17^ Consequently, patients with worse disease severity (and therefore more fluid shifts) may present with a seemingly “larger” vastus measurement, but they actually experience poorer outcomes. These negative correlations thus underscore vastus size as a marker of disease severity at later stages, rather than as a straightforward indicator of healthy muscle mass.

Monitoring changes in vastus size over time may therefore offer important insights into fluid balance and inflammation, helping clinicians gauge the trajectory of illness and recovery potential. ^18^

The rectus femoris demonstrated weaker predictive power compared to the biceps brachii and vastus intermedius. While rectus femoris size correlated with frailty scores, its predictive utility for mortality and post-hospitalization outcomes was limited. This may be due to its susceptibility to atrophy from prolonged immobility rather than systemic illness severity, consistent with findings in prior literature.^19^

Contrary to our initial hypothesis, which predicted a strong correlation between L3SMI and appendicular muscle mass assessed using POCUS, our results revealed no correlation. Additionally, no correlation was found between L3SMI and long-term outcomes. These findings challenge existing assumptions in the field that muscle measurements obtained via POCUS strongly correlate to CT-derived skeletal muscle index.^20^ Several factors may contribute to this unexpected result, such that while L3SMI provides insights into overall core muscle mass, it may not adequately capture peripheral muscle wasting, which is better assessed through direct ultrasound measurements. Peripheral muscle wasting, particularly in weight-bearing muscles like the vastus intermedius, may better reflect clinical trajectories, as evidenced by their stronger correlations with functional outcomes. This discrepancy suggests that different mechanisms underlie core and peripheral muscle atrophy during critical illness, necessitating a nuanced approach to evaluating muscle health and frailty in this population.

This study has several important limitations that should be considered when interpreting our findings. First, it was conducted at a single, tertiary-care academic center, which limits the external validity of our results; patient populations, treatment protocols, and resource availability may differ in smaller community hospitals or centers in other geographic regions. Second, we relied on post-discharge hospital interviews to assess long-term outcomes, and these can be subject to recall bias, incomplete reporting, or loss to follow-up, potentially affecting the accuracy of our outcome measures. Third, while our sample size was sufficient for preliminary analyses, a larger, multicenter cohort would be beneficial to strengthen the statistical power and increase the generalizability of the results. Fourth, muscle thickness measurements are vulnerable to fluid shifts that occur during critical illness, which may confound ultrasound assessments of “true” muscle mass; future work should incorporate robust fluid-balance monitoring or additional imaging modalities to validate muscle changes. Fifth, although we measured muscle size, we did not directly assess functional outcomes such as handgrip strength or walking distance, which may more comprehensively capture the clinical significance of muscle wasting.

## Conclusion

Despite these limitations and some unexpected outcomes, our results open new avenues for investigation, especially regarding the role of localized muscle size assessment with POCUS and its interplay with systemic inflammation, organ dysfunction, and fluid management. Future larger-scale studies that prospectively track muscle function, edema, and inflammatory markers across diverse patient populations will help clarify the mechanisms of peripheral muscle wasting in critical illness and refine our ability to predict patient recovery.

## Methods

### Methodological Approach and Participant Selection

This study was conducted following ethical approval from the Human Studies Protection Office at the Penn State College of Medicine (IRB No. 15328, approved July 30, 2020). All procedures adhered to the ethical guidelines outlined by the Human Studies Protection Office and the Declaration of Helsinki. From April 2023 to July 2024, a Modified Early Warning Score (MEWS)-based algorithm was employed to identify critically ill patients with potential sepsis.^21, 22^ Dual, independent investigator reviews were conducted to ensure unbiased identification of eligible patients from electronically flagged medical records. Informed consent was obtained directly from patients with decision-making capacity or from legally authorized representatives for patients unable to provide consent.

### Inclusion and Exclusion Criteria

Adult patients aged 18 years or older were eligible if identified within 48 hours of critical illness onset and met Sepsis-3 criteria, defined as an increase in SOFA score of two or more in the context of suspected or confirmed infection.^23^ Critical illness was further characterized by the requirement for continuous intravenous vasopressors or noninvasive/invasive respiratory support.

### Clinical Variables and Data Collection

Comprehensive clinical data were obtained through electronic medical records and supplemented by post-discharge interviews where appropriate. Severity of illness was assessed using the Charlson Comorbidity Index^24^ and the Acute Physiology and Chronic Health Evaluation (APACHE) II score.^25^ The primary outcome measure was in-hospital mortality. Secondary outcomes included 30-day Clinical Frailty Score,^26, 27^ 30-day Zubrod/ECOG Performance Status,^28^ 30-and 90-day mortality, 90-day hospital readmission, and number of organ dysfunction-free days. The Zubrod and Clinical Frailty Scores were calculated using a standardized telephone questionnaire that assessed the following criteria: (1) mobility, (2) indicators of assistance with activities of daily living (ADLs), including walking, dressing, transferring, toileting and navigating stairs. Additional adjustments were made based on food intake and weight loss, where severe reductions in food intake or significant weight loss raised the frailty score by one point, emphasizing the impact of nutritional decline.

### Assessment of Low Muscle Mass by Computer Tomography

Axial computed tomography (CT) images, 3.0 mm in thickness, obtained at the L3 vertebral level were retrieved in Digital Imaging and Communication in Medicine (DICOM) format from the Picture Archiving and Communication System (PACS).

Abdominal or pelvic CT scans performed within seven days of the index hospitalization were used to assess skeletal muscle mass.

Skeletal muscle area was manually measured using ImageJ software (version 1.53t; National Institutes of Health, Bethesda, MD, USA),^29^ following standardized protocols.^30, 31^ Both contrast-enhanced and non-contrast CT images were included in the analysis to calculate muscle area, consistent with previously validated methods. Low muscle mass, by CT measurement, was defined as a L3 skeletal muscle index (L3SMI) below the 5th percentile of a sex-matched reference population (41.6 cm²/m² for males and 32.0 cm²/m² for females), as previously reported.^32, 33^

### Peripheral Muscle Measurement by Point-Of-Care Ultrasound

Point-of Care Ultrasound (POCUS) was used to assess the muscle thickness of the biceps brachii, rectus femoris, and vastus intermedius of each patient within seven days of index hospitalization and CT imaging, if applicable. While the patient was lying supine, at 30 degree head elevation and with legs extended and arms supinated, the ultrasound transducer was placed perpendicular to the long axis of the targeted muscle, as previously described.^34, 35^ Briefly, measurement of the biceps brachii muscle was made at the location two-thirds of the distance between the elbow fold and the tip of the acromion.^36^ To measure the rectus femoris and vastus intermedius, the transducer was placed at the location halfway between the anterior superior iliac spine and the patella. All measurements from both the upper and lower extremities were taken from the same laterality. Three consecutive measurements were taken of each muscle by the same investigator on day 1, day 7, and day 14. Muscle measurements were assessed both in raw form and indexed to body surface area, as calculated by the Mosteller formula.^37^

## Statistical Analysis

Our sample size was calculated based on a recent report comparing CT measurements of psoas area with ultrasound measurements of quadriceps depth to discriminate between frail and non-frail patients.^34^ All variables were summarized with descriptive statistics prior to any inferential analysis to assess their distributions. This included using histograms, normal probability plots, and tests for normality for continuous variables. All analyses were performed using SAS version 9.4 (SAS Institute, Cary, NC) using a significance level of 0.05.

A receiver operating characteristic (ROC) analysis utilizing the area under the curve (AUC) was applied to determine the association of the muscle measurements with binary clinical outcomes such as in-hospital mortality, 30-day mortality, 90-day mortality, and 90-day readmission. For each clinical outcome, the Youden Index^38^ was used to find the optimal cut point for each muscle measurement that maximizes both the sensitivity and specificity simultaneously, and the association of this cut point was then tested against the outcome using logistic regression which resulted int an odds ratio to estimate the magnitude and direction of the association.

Spearman correlation was applied to investigate the relationship between the muscle measurements and continuous clinical outcomes. A quantile regression of the median was also used as an alternative approach to determine more specifically the change in the median of the outcome per a 1-unit change in the muscle measurement. Differences in the median of muscle measurements between categories for multinomial clinical outcomes with more than two groups such as discharge location and residence at 30 days were tested with a Kruskal Wallis test including pairwise comparisons using Wilcoxon Rank Sum tests adjusted for multiple group comparisons with the Dwass, Steel, Critchlow-Flinger method^39^.

## Data Availability

All data produced in the present study are available upon reasonable request to the authors

## Acknowledgements

The authors would like to thank Ms. Abigail Samuelsen and Ms. Ruth-Ann Brown for coordinating the study.

## Author Contributions

RA, SDA, and ASB conceptualized and designed the study, while RA and JH developed the study protocol and oversaw data collection. EL performed the statistical analysis and EL, SDA and ASB interpreted the data. RA and ASB drafted the initial manuscript, and JH, SDA, and EL contributed to critical revisions for intellectual content. All authors participated in the final review and approved the manuscript for submission.

## Statements and declarations

## Ethical considerations

Ethical approval was provided by The Penn State College of Medicine Institutional Review Board of the Human Studies Protection Office on Jan 26, 2023 (ref# 15328). All methods were carried out in accordance with relevant institutional guidelines and regulations.

## Consent to participate

Written informed consent was obtained directly from patients with decision-making capacity or from legally authorized representatives (LAR) for patients unable to provide consent. If LARs provided consent over the phone, the name of the LAR, date and time was documented on the informed consent form.

## Data availability

Data are available from the authors on reasonable request.

