## Supplementary Material for "Point-of-Care Ultrasound as a Prognostic Tool in Critically Ill Patients: Insights Beyond Core Muscle Mass"

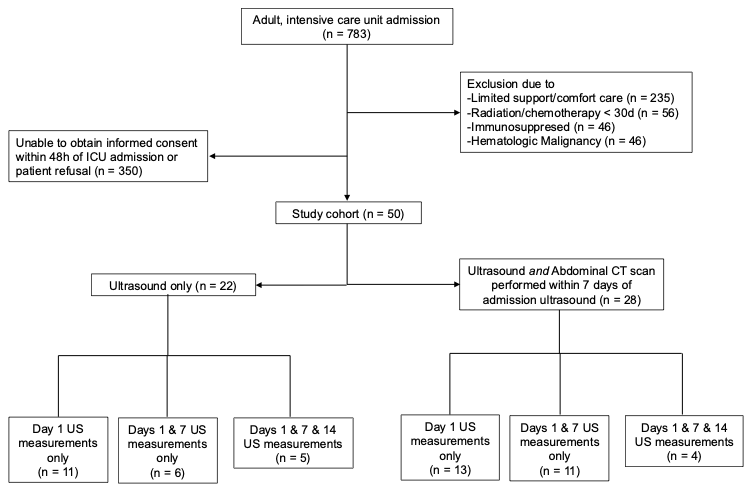

**Fig S1.** STROBE flow chart. STROBE, Strengthening the Reporting of Observational Studies in Epidemiology.
